# Impact of Asymptomatic Enteroaggregative *Escherichia coli* Infection and Co-Pathogen Burden on Intestinal Barrier Function, Linear Growth, and Cognitive Development in Early Childhood: Insights from a Birth Cohort Study

**DOI:** 10.1101/2025.07.04.25330823

**Authors:** Aldo A. M. Lima, Virginia L. S. C. Urtiga, Samilly A. Ribeiro, José Q.S. Filho, Bruna L.L. Maciel, Ana E. S. Alves, Luis F. B. Lino, Francisco A. P. Rodrigues, Ana K. F. Leite, Alexandre Havt, Gagandeep Kang, Margaret N. Kosek, Pascal O. Bessong, Amidou Samie, Rashidul Haque, Estomih R. Mduma, Jose Paulo Leite, Ladaporn Bodhidatta, Najeeha T. Iqbal, Nicola Page, Ireen Kiwelu, Zulfiqar A. Bhutta, Tahmeed Ahmed, Deiziane V S Costa, Richard L. Guerrant, Eric R. Houp

## Abstract

**Introduction:** *Enteroaggregative Escherichia coli* (EAEC) is a major enteric pathogen in low-resource settings and can infect children with or without causing diarrhea. However, limited data exists on the impact of subclinical EAEC infections, particularly when co-detected with other pathogens. This study aimed to evaluate socioeconomic risk factors and the effects of subclinical EAEC infection—alone or in combination with other pathogens—on intestinal barrier function, enteric inflammation, linear growth, and cognitive development in children aged 0–24 months, using quantitative molecular diagnostics.

**Methods:** We analyzed 1,618 non-diarrheal samples from children enrolled in eight sites of the MAL-ED birth cohort. A quantitative PCR-based TaqMan Array Card was used to detect 29 enteropathogens. Children were categorized by infection status: EAEC alone, or EAEC with one, two, or three or more co-pathogens. Associations with socioeconomic indicators (e.g., maternal education, household income, sanitation), clinical symptoms, use medication, anthropometry, biomarkers (myeloperoxidase [MPO], neopterin [NEO], α1-antitrypsin, lactulose:mannitol ratio), nutritional z-scores, and Bayley developmental scores were assessed.

**Results:** Lower socioeconomic and sanitation scores, as well as lower WAMI index values, were significantly associated with EAEC infection and increased pathogen burden (*p < 0*.*001*). Children with subclinical EAEC infection and high pathogen loads also had a higher frequency of antibiotic use. Subclinical EAEC infection—either alone or with ≥3 co-pathogens—was associated with elevated fecal neopterin (β = 687.5; 95% CI: 129.7 to 1245.3), increased intestinal permeability (% lactulose Z-score, β = 0.41; 95% CI: 0.058 to 0.77), lower linear growth (LAZ, β = –0.44; 95% CI: –0.76 to –0.12), and reduced cognitive scores at 15 and 24 months (β = –2.82; 95% CI: –5.15 to –0.48; β = –5.02; 95% CI: –9.81 to –0.22, respectively).

**Conclusion:** Quantitative molecular diagnostics revealed that subclinical EAEC infections with high pathogen burden are associated with adverse socioeconomic conditions, frequent antibiotic use, intestinal inflammation, increased permeability, impaired linear growth, and poorer cognitive development. Even isolated EAEC infection was linked to cognitive deficits, underscoring its potential role in the pathogenesis of environmental enteropathy and early childhood developmental delays.

**What Is Known:**

- Enteroaggregative *Escherichia coli* (EAEC) is a prevalent enteropathogen in children under two years in low-resource settings and has been associated with intestinal inflammation and linear growth faltering.
- Prior studies, largely based on conventional microbiological techniques, reported associations between EAEC and undernutrition, particularly in children with persistent diarrhea or multiple infections.
- The contribution of subclinical EAEC infections—especially in the context of co-infections—to intestinal barrier dysfunction, growth impairment, and cognitive development remained poorly understood.

**What Is New:**

- This study demonstrates that isolated subclinical EAEC infection during the first six months of life is not associated with impaired growth.
- In contrast, EAEC infections co-occurring with three or more pathogens are significantly associated with increased intestinal permeability, elevated fecal neopterin (inflammation), and reduced linear growth (LAZ) in infants.
- Notably, even isolated subclinical EAEC infection is independently associated with lower cognitive and language scores at 15 and 24 months of age.
- These findings, enabled by quantitative molecular diagnostics, underscore the role of EAEC—both alone and in combination with other pathogens—in the pathogenesis of environmental enteric dysfunction and early childhood developmental delays.

## INTRODUCTION

Enteroaggregative *Escherichia coli* (EAEC) is an emerging pathotype characterized by high genetic heterogeneity and is implicated in acute and persistent diarrhea (≥14 consecutive days) in low-resource settings, as well as in diarrheal outbreaks among pediatric and adult populations in high-income countries [1–3]. Furthermore, EAEC has been associated with traveler’s diarrhea [4] and diarrhea in individuals with human immunodeficiency virus (HIV) [4, 5]. Beyond its well-documented role in diarrheal morbidity, EAEC infection has been implicated in asymptomatic colonization, undernutrition [6], and co-infection with multiple enteropathogens, particularly in cases involving three or more added pathogens, which has been associated with impaired linear growth [7].

In the Malnutrition and Enteric Diseases (MAL-ED) birth cohort study, EAEC was identified as the second most prevalent enteropathogen in both diarrheic and non-diarrheic surveillance stool samples from children aged 0 to 11 months and the third most prevalent pathogen in children aged 12 to 24 months [2]. However, subsequent reanalysis utilizing molecular diagnostics revealed a substantial reduction in EAEC prevalence, ranking it as the 11th most frequently detected enteropathogen among children with and without diarrhea across the first two years of life [8]. Despite this reduction in ranking, EAEC stayed a pathogen of significant clinical and epidemiological relevance during early childhood.

EAEC infection has also been linked to intestinal inflammation and compromised barrier function in children showing growth faltering, underscoring its potential role in the pathogenesis of environmental enteropathy in resource-limited settings [9, 10]. An earlier study by our group evaluated the impact of asymptomatic EAEC detection and co-infections in children aged 0 to 6 months from the MAL-ED cohort using conventional microbiological methods and multiplex polymerase chain reaction assay for enteropathogen detection [7]. This study showed an association between EAEC infection, particularly in the presence of three or more co-infecting pathogens, and increased intestinal inflammation, dysregulated immune responses, and malnutrition [7].

Given the superior sensitivity and specificity of molecular diagnostics compared to conventional culture-based methods [8], the present study aims to comprehensively reassess the risk factors, intestinal integrity, inflammatory markers, and innate immune responses associated with EAEC infection, both as a monoinfection and in the context of enteropathogen co-infections, using a quantitative molecular diagnostic approach. Furthermore, we will evaluate the impact of these infections on early-life growth outcomes during the first six months of life across all eight MAL-ED study sites. Our central hypothesis is that molecular quantification of enteropathogen burden will provide enhanced resolution in characterizing population-level determinants of EAEC-associated infections and their synergistic interactions with other enteropathogens, thereby facilitating a more precise understanding of the consequences of subclinical EAEC enteric infection and co-infections on intestinal barrier function, inflammation, early childhood growth trajectories and cognitive development.

## MATERIALS AND METHODS

### Population, ethics and study design

This study utilized data from children aged 0 to 24 months from the multicenter MAL-ED project, which followed children from birth to 5 years of age to investigate etiology, risk factors, and interactions between enteric infections and malnutrition across eight developing countries. The study sites included Dhaka, Bangladesh; Fortaleza, Brazil; Vellore, India; Bhaktapur, Nepal; Naushahro Feroze, Pakistan; Loreto, Peru; Venda, South Africa; and Haydom, Tanzania. Ethical approval was granted by the National Research Ethics Commission (CONEP) (approval No. 029/10) and the Research Ethics Committee of the Federal University of Ceará (COMEPE/UFC) (protocol No. 246/09). Informed consent was obtained from the parents or legal guardians of each child. A total of 2,145 newborns, up to 17 days old, were enrolled between November 2009 and February 2012. We employed an unpaired nested case-control design to minimize selection, information, and reverse causality biases. The overall study design has been described in detail in [11, 12]. Study enrollment flowchart can be seen in **Figure Suppl. 1** (**Supplementary Material**).

### Surveillance, stool collection and diagnosis of enteropathogens

Surveillance was carried out during twice-weekly home visits. Caregivers responded to a standardized questionnaire designed to collect daily symptoms data, including cough, fever, vomiting, diarrhea, and use of medication during these visits [11]. We utilized non-diarrheal stool samples collected during surveillance. The detection of 29 enteropathogens was performed using the quantitative molecular TAC array analysis (Thermo Fisher, Carlsbad, CA, USA), as previously described [8, 13]. All procedures, including sample storage, nucleic acid extraction, assay validation, quantitative PCR setup, and quality control, are detailed in Platts-Mills et al. [8]. The genes *aatA* and/or *aaiC* were used as criteria for the molecular diagnosis of EAEC in stool samples.

### Socioeconomic status and maternal education

Structured questionnaires were administered to collect information on birth date, sex, anthropometric data, childcare practices, maternal or caregiver characteristics, household composition, maternal education, household sanitation conditions, household assets, and average monthly income. Socioeconomic and sanitation (SES) status were assessed based on family income, maternal education, access to potable water, sanitation infrastructure, and household assets (measured through an asset score including eight household items). The WAMI index, which evaluates access to water and sanitation, asset ownership, maternal education level, and household income, was also applied. During weekly visits, caregivers also completed a standardized questionnaire to collect records on basic dietary intake (breastfeeding). The scoring system for these and other categorical variables is described in LIMA et al. [7]. Clinical data, breastfeeding practices, and medication use were analyzed in terms of the percentage of days of occurrence, enabling an assessment of children’s health conditions.

### Anthropometric measurements

A standard recumbent length board (Schorr Productions, Olney, MD) was utilized to measure the monthly length of all enrolled children with an accuracy of 0.1 cm [7]. Digital scales with a precision of 100 g were used for monthly weight measurements. The World Health Organization (WHO) Multicenter Growth Reference Study was employed to calculate z-scores for weight-for-age (WAZ), length-for-age (LAZ), and weight-for-length (WLZ) [14]. Anthropometric data from Pakistan were excluded due to concerns about measurement quality.

### Intestinal integrity, immune and inflammatory response

The lactulose and mannitol test were employed to assess intestinal permeability, absorptive ability, and mucosal damage following a previously standardized protocol [15]. This test was administered to children at 3 and 6 months of age, with the meaning of these two measurements used for analysis. Additionally, three fecal biomarkers were evaluated in non-diarrheic stool samples collected monthly between birth and 6 months of age, using a standardized protocol and specific data collection tools [15, 16]. The biomarkers evaluated included alpha-1-antitrypsin (A1AT), whose elevated levels show severe intestinal barrier damage; myeloperoxidase (MPO), a marker of intestinal inflammation; and neopterin (NEO), which, when present at high concentrations, is associated with enhanced mucosal immune activation [3]. Median values of these biomarkers were used for the present analysis.

**Child neurodevelopment**

The Bayley Scales of Infant and Toddler Development, Third Edition (BSID-III) [17] were used to assess cognitive development in children at 6, 15, and 24 months of age across the MAL-ED study sites. The present analysis aimed to evaluate the association between early-life subclinical EAEC enteric infections, co-pathogen subclinical infections, and subsequent neurodevelopment. Trained assessors, with expertise in psychology or child development, administered the BSID-III in standardized settings—quiet, well-lit, central locations—on days when the children were healthy.

### Statistical analysis

Children were included in the analysis if they had at least 90% adherence to the surveillance protocol, completed the socioeconomic follow-up form, and had molecular diagnostic results for EAEC and 28 other enteropathogens across monthly stool samples collected during the first six months of life. Based on cumulative infection profiles, participants were categorized into seven mutually exclusive groups: 1. No pathogen detected; 2. EAEC only; 3. EAEC plus one co-pathogen; 4. EAEC plus two co-pathogens; 5. EAEC plus three or more co-pathogens; 6. One or two non-EAEC pathogens; 7. Three or more non-EAEC pathogens. Descriptive statistics were employed to summarize categorical variables (e.g., maternal education, sanitation status, water source) and continuous variables (e.g., monthly income, anthropometric indices, biomarker levels). Comparisons between groups were performed using one-way analysis of variance (ANOVA) or Kruskal-Wallis tests for continuous variables, and chi-square or Fisher’s exact tests for categorical variables, depending on data distribution and sample size. Post-hoc pairwise comparisons with appropriate corrections (e.g., Tukey or Dunn’s tests) were conducted when omnibus tests indicated significant differences. To assess associations between infection group and outcomes related to intestinal permeability, inflammatory markers (e.g., MPO, A1AT, NEO), and anthropometric measures (WAZ, LAZ, WLZ), generalized linear models and multivariable linear regression analyses were applied. The models were adjusted for potential confounders identified as significant in the univariate analysis, including study location, household food insecurity, maternal education, socioeconomic status (WAMI index), and the percentage of days with antibiotic use. All statistical analyses were conducted using SPSS version 25.0 (IBM Corp., Armonk, NY, USA), GraphPad Prism® version 8.0.2 (GraphPad Software, Boston, MA, USA), and Jamovi version 2.6. Summary figures, including the abstract schematic and flow diagrams, were created using Canva (Canva Pty Ltd., Sydney, Australia). Statistical significance was defined as a two-tailed p-value < 0.05.

## RESULTS

### Selection of the study population and association of potential risk determinants for isolated or combined subclinical enteroaggregative *Escherichia coli* infections

Of the 2,145 children enrolled in the cohort between November 2009 and February 2012, a total of 1,618 (75.43%) were selected based on at least 90% active surveillance follow-up until six months of age. **Table 1** summarizes the categorical risk variables associated with subclinical Enteroaggregative *Escherichia coli* (EAEC) infections, either in isolation or in combination with other enteropathogens, during the first six months of life. We identified that there was a significant difference between the groups (*p < 0*.*001*) in relation to the study location, however we did not identify a significant difference in relation to sex and birth weight <2,500g. Maternal education <6 years, inadequate sanitation, and unsafe drinking water sources were significantly more prevalent among children with asymptomatic EAEC detections co-occurring with ≥3 additional pathogens compared to all other groups (*p = 0*.*007*; *p < 0*.*001*; *p < 0*.*001*, respectively). Conversely, food insecurity was significantly higher in the group with ≥3 pathogens excluding EAEC (*p = 0*.*001*).

**Table 1.** Determinant Categorical Variables Associated with the Presence of EAEC on monthly samples without diarrhea from 0 to 6 Months of Age.

|  |  | 1 or 2<br>Pathogens<br>Without<br>EAEC | 3 or More<br>Pathogens<br>Without EAEC | EAEC_P0 | EAEC_P1 | EAEC_P2 | EAEC_P3 | No<br>Pathogens<br>Detected | p |
| --- | --- | --- | --- | --- | --- | --- | --- | --- | --- |
| **Location (%)** | BG | 3 (3.3%) | 23 (14.6%) | 0 (0%) | 5 (7.2%) | 7 (7%) | 167 (14.4%) | 0 (0%) | **<0,001\*** |
|  | BR | 28 (30.8%)* | 27 (17.1%)* | 8 (25.8%)* | 12 (17.4%) | 16 (16%) | 53 (4.6%) | 21 (40.4%)* |  |
|  | IN | 4 (4.4%) | 5 (3.2%) | 1 (3.2%) | 6 (8.7%) | 5 (5%) | 205 (17.7%)* | 0 (0%) |  |
|  | NP | 32 (35.2%)* | 31 (19.6%)* | 9 (29%)* | 15 (21.7%)* | 21 (21%)* | 104 (9%) | 15 (28.8%)* |  |
|  | PE | 2 (2.2%) | 18 (11.4%) | 3 (9.7%) | 4 (5.8%) | 13 (13%) | 144 (12.4%) | 3 (5.8%) |  |
|  | PK | 8 (8.8%) | 24 (15.2%) | 6 (19.4%) | 13 (18.8%)* | 11 (11%) | 158 (13.6%) | 7 (13.5%) |  |
|  | SA | 14 (15.4%) | 27 (17.1%)* | 4 (12.9%) | 11 (15.9%) | 23 (23%)* | 135 (11.7%) | 5 (9.6%) |  |
|  | TZ | 0 (0%) | 3 (1.9%) | 0 (0%) | 3 (4.3%) | 4 (4%) | 192 (16.6%)* | 1 (1.9%) |  |
|  | Total n (%) | 91 (100%) | 158 (100%) | 31 (100%) | 69 (100%) | 100 (100%) | 1158 (100%) | 52 (100%) |  |
| **Female / Male** | **(male)** | 46 (50.5%) | 84 (53.2%) | 11 (35.5%) | 39 (56.5%) | 53 (53%) | 591 (51%) | 24 (46.2%) | 0,575 |
| **n (%)** | Total n (%) | 91 (100%) | 158 (100%) | 31 (100%) | 69 (100%) | 100 (100%) | 1158 (100%) | 52 (100%) |  |
| **Birth Weight <2500g** | **(yes)** | 9 (10.7%) | 15 (10.9%) | 2 (7.4%) | 6 (10.9%) | 8 (9.2%) | 103 (11%) | 2 (4.4%) | 0,877 |
| **n (%)** | Total n (%) | 84 (100%) | 138 (100%) | 27 (100%) | 55 (100%) | 87 (100%) | 935 (100%) | 45 (100%) |  |
| **Mother with < 6 Years of Education** | **(yes)** | 22 (25%) | 58 (38.9%) | 9 (32.1%) | 21 (33.3%) | 24 (26.4%) | 440 (39.6%)* | 12 (24%) | **0,007\*** |
| **n (%)** | Total n (%) | 88 (100%) | 149 (100%) | 28 (100%) | 63 (100%) | 91 (100%) | 1110 (100%) | 50 (24%) |  |
| **Source of Potable Water** | **(Inadequate)** | 2 (2.3%) | 5 (3.4%) | 1 (3.6%) | 4 (6.3%) | 5 (5.5%) | 150 (13.5%)* | 1 (2%) | **<0,001\*** |
| **n (%)** | Total n (%) | 88 (100%) | 149 (100%) | 28 (100%) | 63 (100%) | 91 (100%) | 1111 (100%) | 50 (100%) |  |
| **Sanitationn (%)** | **(Inadequate)** | 5 (5.7%) | 26 (17.4%) | 4 (14.3%) | 12 (19%) | 18 (19.8%) | 426 (38.3%)* | 5 (10%) | **<0,001\*** |
|  | Total n (%) | 88 (100%) | 149 (100%) | 28 (100%) | 63 (100%) | 91 (100%) | 1112 (100%) | 50 (100%) |  |
| **Food Insecurity** | **(some)** | 39 (51.3%) | 80 (56.7%)* | 16 (61.5%) | 32 (50%) | 46 (53.5%) | 455 (42.4%) | 30 (62.5%) | **0,001\*** |
| **n (%)** | Total n (%) | 76 (100%) | 141 (100%) | 26 (100%) | 64 (100%) | 86 (100%) | 1074 (100%) | 48 (100%) |  |

Each subset of groups of children whose column proportions significantly differ from each other at the 0.05 level were reported, with residual values above 3 identified among the investigated groups.
Enteraggregative *Escherichia coli* (EAEC) with or none (P0), 1 (P1), 2 (P2), and 3 (P3) or more any other pathogens.
Location: BG, Bangladesh; BR, Brazil; IN, India; NP, Nepal; PE, Peru; PK, Pakistan; SA, South Africa; and TZ, Tanzania.
\* Shows a statistically significant difference in the variable between groups.

**Table 2** presents the quantitative determinant variables associated with EAEC infection during the first six months of life. No significant difference in birth weight was seen between the investigated groups (*p > 0*.*3*). However, monthly income was significantly lower (*p < 0*.*00*1) in children with EAEC and ≥ 3 pathogens compared to all other groups. Monthly income was also lower in the EAEC with 2 other pathogens group when compared to children with 1 or 2 pathogens without EAEC (*p = 0*.*0266*) and pathogen-free children (*p = 0*.*0325*). Mother’s years of education were significantly lower in the EAEC with ≥3 more pathogens group compared to the groups: ≥3 pathogens without EAEC (*p = 0*.*0012*), one or two pathogens without EAEC (*p < 0*.*000*1), EAEC with one more pathogen (*p < 0*.*0001*) and pathogen-free (*p = 0*.*0087*). Mother’s education was also lower in children with EAEC and two pathogens when compared to children with one or two pathogens without EAEC (*p = 0*.*0226*) and children with EAEC and one additional pathogen (*p = 0*.*0277*). SES, measured using asset-based scoring, was lower in the EAEC with ≥3 other pathogens group compared to all other groups (*p < 0*.*001*) and in the ≥3 pathogens without EAEC group when compared to the pathogen-free group. Furthermore, we also identified lower SES in the EAEC and 2 other pathogens and ≥3 pathogens without EAEC groups when compared to children with 1 or 2 pathogens without EAEC (*p = 0*.*0085, p = 0*.*0208*, respectively). A similar pattern was observed for the WAMI index (water/sanitation, household assets, maternal education, and household income), with the EAEC with ≥3 additional pathogens group having the lowest scores (*p < 0*.*001*) compared with all other groups. The WAMI index was also significantly lower in the EAEC, and two other pathogens group compared with the one or two pathogens without EAEC and no pathogens groups (*p = 0*.*0005* and *p = 0*.*0152*, respectively). The proportion of days with the use of antibiotics was significantly higher in children with EAEC combined with ≥3 additional pathogens and children with 3 or more pathogens without EAEC compared with children with one or two pathogens without EAEC and children without pathogens (*p ≤ 0*.*001*). We also identified that children with EAEC and one or two other pathogens had higher use of antibiotics compared with children with two or more pathogens without EAEC (p = 0.0004, p = 0.0454, respectively), as well as children with EAEC and two other pathogens had higher use of antibiotic than children without pathogens (*p = 0*.*0025*). In contrast, the proportion of days with acute respiratory infection, breastfeeding, dehydration, diarrhea, and fever was similar between the investigated groups (*p > 0*.*1*).

**Table 2.** Determinant Quantitative Variables Associated with the Presence of Enteroaggregative *Escherichia coli* on monthly samples without diarrhea from 0 to 6 Months of Age.

|  | 1 or 2 Pathogens<br>Without EAEC (A) | 3 or More<br>Pathogens<br>Without EAEC<br>(B) | EAEC_P0 (C) | EAEC_P1 (D) | EAEC_P2 (E) | EAEC_P3 (F) | No Pathogens<br>Detected (G) | p | DIFFERENCES |
| --- | --- | --- | --- | --- | --- | --- | --- | --- | --- |
| **Birth Weight (kg)**<br>**Median (P25 – P75)** | 3.11<br>(2.75 - 3.42) | 2.97<br>(2.64 - 3.31) | 3.18<br>(2.8 - 3.5) | 3.1<br>(2.8 - 3.5) | 3.<br>(2.75 - 3.3) | 3<br>(2.77 - 3.3) | 3.1<br>(2.95 - 3.39) | > 0,3 | - |
| **Monthly Income (US\$)**<br>**Median (P25 – P75)** | 190.4<br>(108.8 – 346.7) | 190.4<br>(108.8 – 346.7) | 150.8<br>(94.15 – 245.1) | 164.3<br>(95.03 – 298.4) | 132**<br>(83.8 - 230) | 98*<br>(54.3 – 162.7) | 232.0<br>(117 - 365.8) | <0,001 | FT*<br>EB, EG*** |
| **Mother's Years of Education**<br>**Median (P25 – P75)** | 9<br>(5.5 - 11) | 9<br>(5 - 11) | 9<br>(5 - 12) | 9<br>(5 - 12) | 7***<br>(3 - 10) | 7*<br>(3 - 9) | 9<br>(5 - 11) | <0,001 | FA, FB, FD,<br>FG*<br>EA, ED*** |
| **SES (sum of 8 asset scores)**<br>**Median (P25 – P75)** | 6<br>(5 - 7) | 5**<br>(3 - 6) | 6<br>(4.25 - 7) | 6<br>(3 - 7) | 5***<br>(3 - 7) | 4*<br>(2 - 6) | 7<br>(6 - 8) | <0,05 | FT*<br>BA, BG**<br>EA, EG*** |
| **WAMI Index**<br>**Median (P25 – P75)** | 0.76<br>(0.6 - 0.85) | 0.64**<br>(0.48 - 0.81) | 0.69<br>(0.56 - 0.84) | 0.68<br>(0.56 - 0.82) | 0.6<br>(0.48 - 0.75) | 0.5*<br>(0.34 - 0.64) | 0.75**<br>(0.60 - 0.85) | <0,05 | FT*<br>EA, EG*** |
| **% of Days with Lower**<br>**Respiratory Tract Illness**<br>**Median (P25 – P75)** | 1.15<br>(0 - 4.87) | 2.7<br>(1.1 - 3.4) | 1.65<br>(0 – 3.67) | 2.5<br>(0 - 4.32) | 0.5<br>(0 – 2.95) | 0<br>(0 – 2.9) | 0.55<br>(0 – 3.92) | 0,661 | - |
| **% of Days with Antibiotic**<br>**Median (P25 – P75)** | 0<br>(0 - 4.07) | 4**<br>(0 - 11) | 1.6<br>(0 – 9.95) | 2.7<br>(0 – 10.6) | 3.8***<br>(0 – 10.5) | 3.8*<br>(0 - 10.3) | 0<br>(0 - 4.3) | <0,001 | FA, FG*<br>BA, BG**,<br>EB, EG***<br>AD# |
| **% of Days with**<br>**Breastfeeding**<br>**Median (P25 – P75)** | 98.3<br>(95.1 - 99.75) | 98.4<br>(96.7 - 98.9) | 97.7<br>(93.5 - 98.9) | 97.7<br>(94.5 - 92.28) | 97.8<br>(94.1 - 98.9) | 98.3<br>(96.6 - 98.9) | 98.3<br>(93 - 100) | >0,9 | - |
| **% of Days with**<br>**Dehydration**<br>**Median (P25 – P75)** | 4.4<br>(2.7 - 36.4) | 3.9<br>(1.95 - 8.9) | 9.2<br>(4.65 - 13.65) | 2,75<br>(1.32 – 5.95) | 4.3<br>(2.8 – 8.7) | 4.9<br>(2.7 – 9.6) | 5.9<br>(4.05 - 7.60) | >0,99 | - |
| **% of Days with Diarrhea**<br>**Median (P25 – P75)** | 0<br>(0 - 3.2) | 1.6<br>(0 - 6.25) | 0<br>(0 – 4.1) | 1.6<br>(0 - 5.45) | 1.6<br>(0 - 5.5) | 1.1<br>(0 - 4.9) | 0<br>(0 - 3.4) | >0,1 | - |
| **% of Days with Fever**<br>**Median (P25 – P75)** | 0<br>(0 - 0) | 0<br>(0 - 0) | 0<br>(0 - 0) | 0<br>(0 - 0) | 0<br>(0 - 0) | 0<br>(0 - 0) | 0<br>(0 - 0) | >0,99 | - |

*Escherichia coli* enteroaggregative (EAEC) with or none (P0), 1 (P1), 2 (P2), and 3 (P3) or more any other pathogens. The groups were named by letters (A, B, C, D, E, F, G), and each subset of groups of categories of children whose column proportions significantly differ from each other at the 0.05 level was reported in the "differences" column. The letter "T" refers to all study groups.
Results are presented as median with interquartile range (25th–75th percentile).
The statistical analysis used was the Kruskal-Wallis tests for all groups, $p < 0.05$ .
The symbols in the table mean statistical difference between:
\* EAEC with 3 or more pathogens vs. other groups specified in the table.
\*\* 3 or more pathogens without EAEC vs. other groups specified in the table.
\*\*\* EAEC and 2 other pathogens vs. other groups specified in the table.

### Influence of subclinical infection by EAEC and copathogens on markers of gastrointestinal functional and immunoinflammatory responses

A higher pathogen burden, both in the presence and absence of EAEC, was significantly associated with increased Z-score–corrected relative lactulose excretion compared to children without any detected pathogens, but only after adjustment for covariates (EAEC_P3: β = 0.41; 95% CI: 0.06 to 0.77; No EAEC_P3: β = 0.44; 95% CI: 0.047 to 0.84) (**Figures 1A1** and **1A’1**). Elevated myeloperoxidase levels were significantly associated with children harboring EAEC and three or more pathogens, relative to pathogen-free children, in the unadjusted model (EAEC_P3: β = 3509.4; 95% CI: 788 to 6231); however, this association lost significance after adjusting for covariates (EAEC_P3: β = 693.8; 95% CI: –2069.8 to 3457.4) (**Figures 1B1** and **1B’1**). In contrast, increased neopterin levels were significantly associated with the presence of EAEC plus one or three additional pathogens only in the adjusted models (EAEC_P1: β = 1016.4; 95% CI: 314.4 to 1718.5; EAEC_P3: β = 687.5; 95% CI: 129.7 to 1245.3) (**Figures 1B2** and **1B’2**).

**Figure 1.**
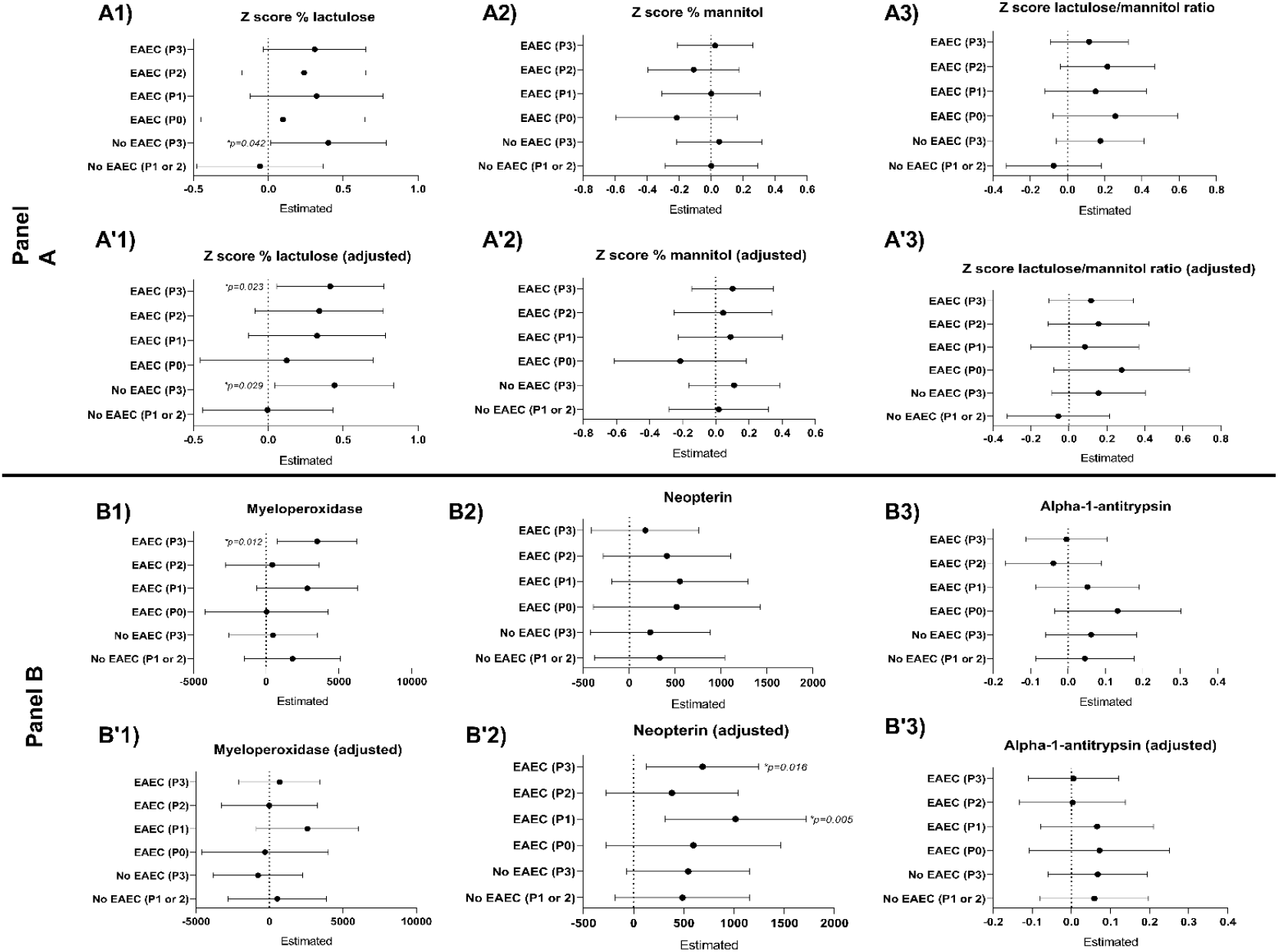
Unadjusted and adjusted linear regression models were used to evaluate the association between asymptomatic EAEC detection (with or without copathogens) and the following outcomes in children aged 0 to 6 months. Panel A, Z-score–corrected % lactulose excretion (A1 no adjusted and A’1 adjusted), % mannitol excretion (A2 no adjusted and A’2 adjusted), lactulose:mannitol ratio (A3 no adjusted and A’3 adjusted). Panel B, myeloperoxidase (ng/mL; B1 no adjusted and B’1 adjusted), neopterin (nmol/L; B2 no adjusted and B’2 adjusted), and alpha-1-antitrypsin (mg/g; B3 no adjusted and B’3 adjusted). All biomarker values were summarized using medians. Z-scores for % lactulose, % mannitol, and the lactulose:mannitol ratio were calculated based on the median of values measured at 3 and 6 months to reflect the cumulative interval from 0 to 6 months. Myeloperoxidase, neopterin, and alpha-1-antitrypsin values also reflect median levels across the cumulative 0–6-month interval. The groups are enteroaggregative *Escherichia coli* (EAEC) with or none (P0), 1 (P1), 2 (P2), and 3 (P3) or any other pathogens. All groups were compared to children with no detected enteric pathogens. Adjusted models included the following covariates: study site, household food insecurity, maternal education, socioeconomic status (WAMI index), and percentage of days with antibiotic use. Results are presented as β estimates with 95% confidence intervals.

No significant associations were found for Z-score–corrected relative mannitol excretion, lactulose/mannitol ratio, or alpha-1-antitrypsin in either adjusted or unadjusted models (**Figures 1A2, 1A’2, 1A3, 1A’3, 1B3, 1B’3**). For detailed regression coefficients, confidence intervals, and statistical difference refer to **Table 1** in the **supplementary material**.

### Association of asymptomatic EAEC detections, isolated or combined with any other pathogen with undernutrition

Regarding longitudinal anthropometric outcomes, a higher pathogen burden—both in the absence of EAEC and in the presence of EAEC with 1, 2, or ≥3 co-infecting pathogens—was significantly associated with a decline in length-for-age z-score (LAZ delta) compared to pathogen-free children in the unadjusted model (**Figure 2A**). However, in the adjusted model, only children with EAEC and ≥3 pathogens showed a significant association with decreased LAZ delta (EAEC_P3: adjusted β = –0.44; 95% CI: –0.76 to –0.12) (**Figure 2B**). Similarly, children with EAEC and ≥3 pathogens had significantly lower weight-for-age z-scores (WAZ delta) in the unadjusted model compared to pathogen-free children, although this association was no longer significant after adjustment (**Supplement Table 2**). In contrast, weight-for-length z-scores (WLZ delta) were consistently higher among children with EAEC and two more pathogens in both unadjusted and adjusted models (**Supplement Table 2**).

**Figure 2.**
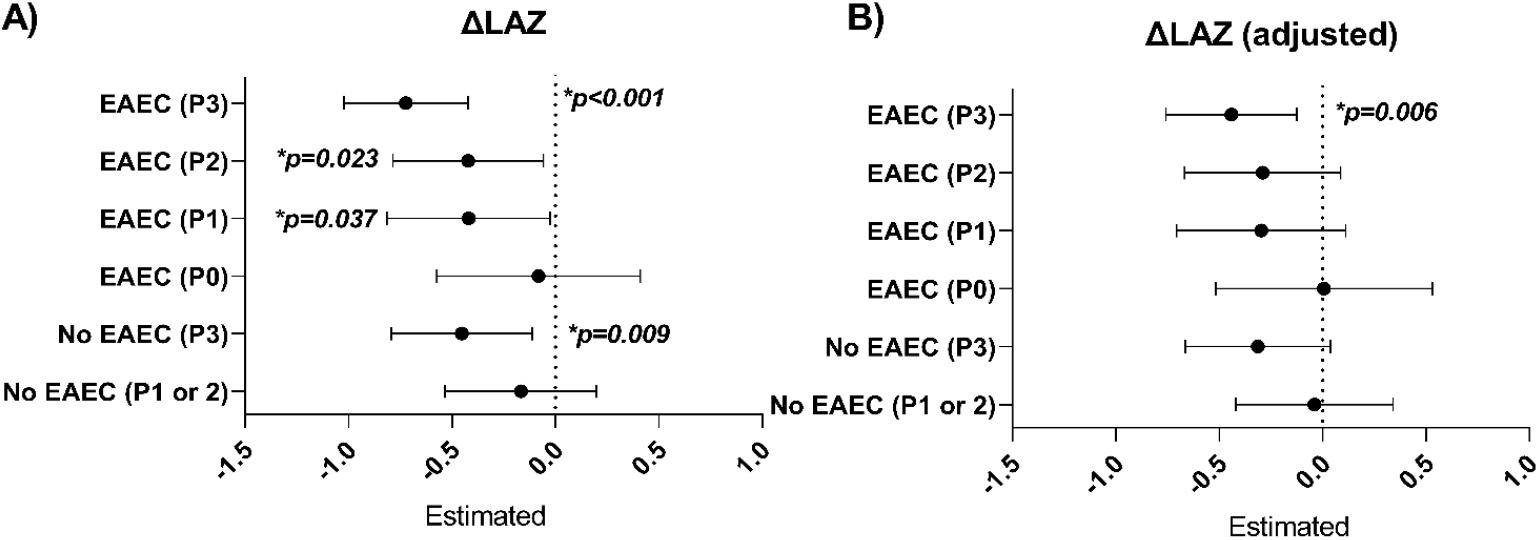
Unadjusted and adjusted linear regression models were used to assess changes in delta length-for-age Z-score (ΔLAZ), A) unadjusted and B) adjusted, of children 0–6 months of age, with or without subclinical EAEC and copathogen infection. The groups are enteroaggregative Escherichia coli (EAEC) with or none (P0), 1 (P1), 2 (P2), and 3 (P3) or any other pathogens. All groups were compared to children with no detected pathogens. Deltas were calculated using anthropometric values at 0 and 6 months. Adjusted models included the following covariates: study site, household food insecurity, maternal education, socioeconomic status (WAMI index), and percentage of days with antibiotic use. Results are presented as β estimates with 95% confidence intervals.

### Association of asymptomatic EAEC detections, isolated or combined with any other pathogen with child neurodevelopment

Neurodevelopmental outcomes at 6 months of age did not significantly predict BSID-III composite cognitive scores (data not shown). **Figure 3** and **Supplementary Table 3** presents the results of unadjusted and adjusted linear regression models assessing the association between early-life subclinical infection and neurodevelopmental outcomes at 15 and 24 months, as measured by the Bayley Scales of Infant and Toddler Development (BSID-III). After adjusting for potential confounders—including study site, household food insecurity, maternal education, socioeconomic status (WAMI index), and antibiotic use—children with early subclinical infection by *Enteroaggregative Escherichia coli* (EAEC) alone (P0 group) showed significantly lower BSID-III composite cognitive scores at both 15 months (β = −2.82; 95% CI: −5.15, −0.48; *p = 0*.*018*) and 24 months (β = −5.02; 95% CI: −9.81, −0.22; *p = 0*.*040*), compared to children without any pathogen detection (**Figure 3A’1** and **3B’1**). Additionally, this same group showed a significant reduction in total language scores at 24 months (β = −6.92; 95% CI: −13.38, −0.47; *p = 0*.*036*, **Figure 3B’2**). No other domains, including total motor and social-emotional scores, showed statistically significant associations in any exposure group after adjustment. These findings suggest that early subclinical EAEC mono-infection may have a specific and lasting detrimental impact on cognitive and language development during early childhood.

**Figure 3.**
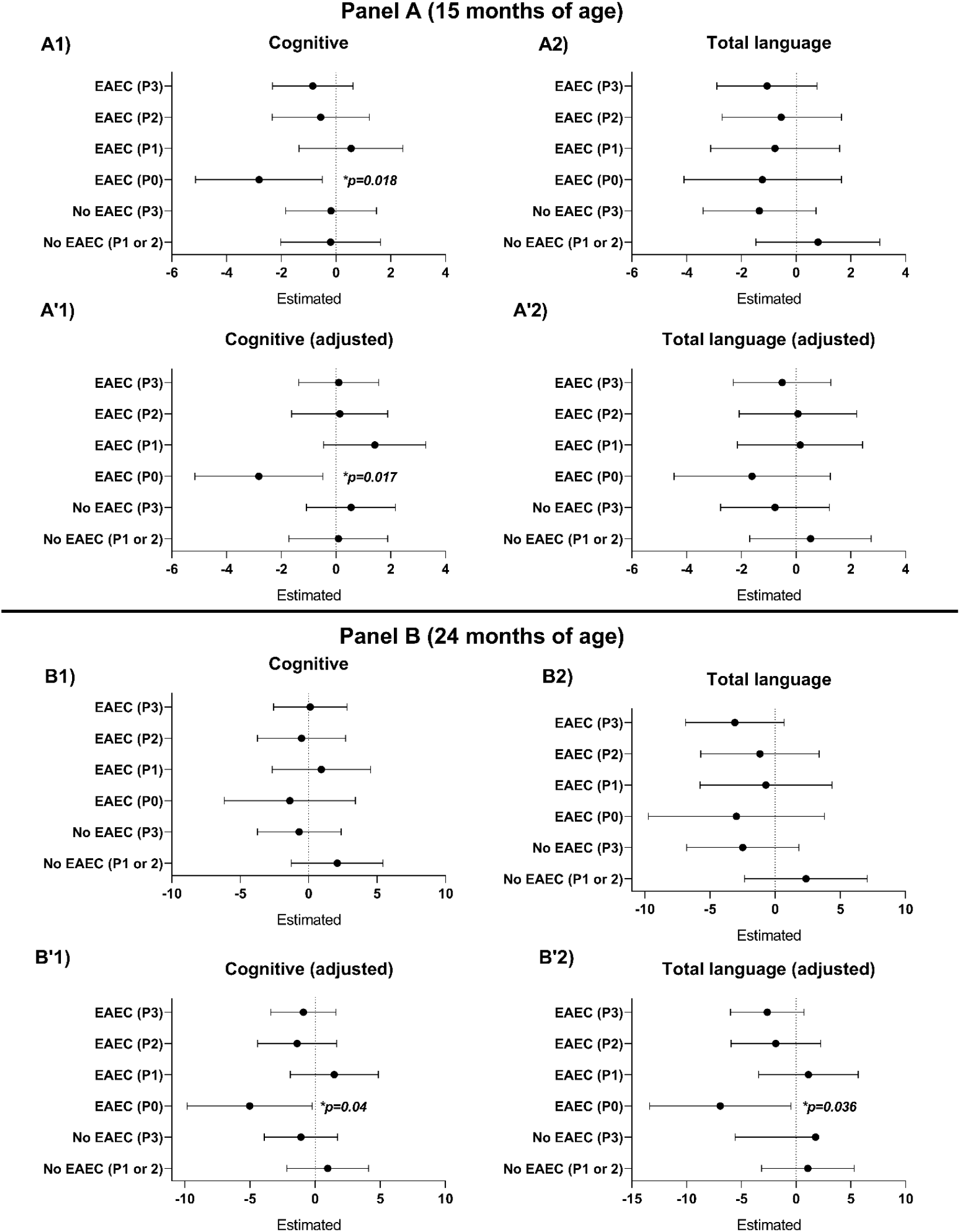
General linear models (unadjusted and adjusted) were used to assess Bayley cognitive and total language scores in children aged 15 and 24 months with or without subclinical infection. Panel A presents data for children aged 15 months: A1) unadjusted cognitive scores, A’1) adjusted cognitive scores, A2) unadjusted total language scores, and A’2) adjusted total language scores. Panel B presents corresponding data for children aged 24 months: B1) unadjusted cognitive scores, B’1) adjusted cognitive scores, B2) unadjusted total language scores, and B’2) adjusted total language scores. Groups were categorized according to the presence of enteroaggregative Escherichia coli (EAEC) alone (P0), or in combination with one (P1), two (P2), or three (P3) additional enteropathogens. All groups were compared to children with no detected pathogens. Adjusted models included study site, household food insecurity, maternal education, socioeconomic status (WAMI index), and percentage of days with antibiotic use. Results are shown as β estimates with 95% confidence intervals.

### Prevalence of subclinical enteric co-infections associated with EAEC

Given that children with EAEC co-infected with two or three or more additional pathogens showed notable associations with nutritional status and biomarkers of intestinal function and immune activation, we further investigated which pathogens were co-detected in these groups. Among children with EAEC and two more pathogens, four pathogens were shown in more than 20% of samples: atypical enteropathogenic *Escherichia coli* (aEPEC), enterotoxigenic *E. coli* (ETEC), adenovirus 40/41, and norovirus genogroups GI or GII (**Figure 4A**). In contrast, children with EAEC and three or more co-infecting pathogens showed >20% prevalence for ten pathogens: ETEC, aEPEC, norovirus genogroups GI or GII, adenovirus 40/41, astrovirus, Campylobacter jejuni or C. coli, typical EPEC (tEPEC), Giardia, sapovirus, and rotavirus (**Figure 4B**).

**Figure 4.**
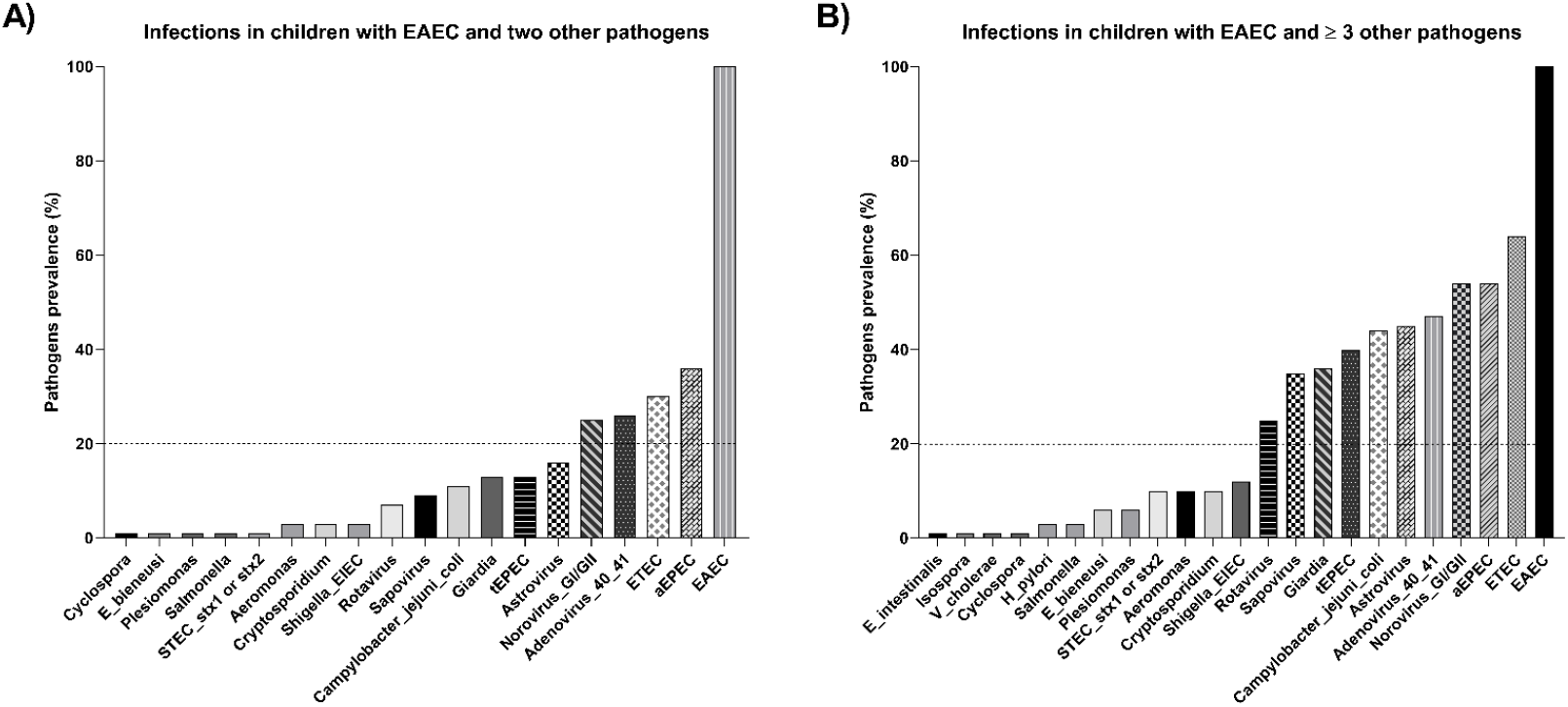
Prevalence of subclinical coinfection in children with EAEC and (A) two or (B) ≥ 3 other pathogens in stool samples from children in the first six months of life. EAEC = enteroaggregative *Escherichia coli*; aEPEC = atypical enteropathogenic *E coli*; tEPEC = typical enteropathogenic *E coli*; tEPEC = typical enteropathogenic *E coli*; ETEC = enterotoxigenic *E coli*; *Shigella*/EIEC = *Shigella* or Enteroinvasive *E. coli*; STEC = Shiga toxin-producing *Escherichia coli* stx1 or stx2; *Campylobacter_jejuni_coli* = C. jejuni or C. coli; *E_bieneusi* = *Enterocytozoon bieneusi*; *E_intestinalis* = *Enterocytozoon intestinalis*. Pathogens present in <1% of stool samples are not shown.

## DISCUSSION

Enteroaggregative *Escherichia coli* (EAEC) has been recognized as a significant enteric bacterial pathogen in low-income settings, particularly affecting children under two years of age—both with and without overt diarrhea [9, 18, 19]. While several studies have shown associations between EAEC infection and various risk factors [20, 21], few have examined its role in subclinical infections [9] or its co-occurrence with multiple enteropathogens [7]. Moreover, evidence is still limited about the relationship between EAEC infection, intestinal inflammation, and linear growth faltering during early childhood [9, 10]. Within this context, the present study assessed the impact of EAEC infection—both in isolation and in conjunction with other pathogens—on linear growth, intestinal barrier function, and enteric inflammation. The main finding was that asymptomatic EAEC detection in early childhood was associated with increased intestinal paracellular permeability, activation of the intestinal immune response, impaired linear growth, and poorer cognitive development.

Co-infections in non-diarrheic stool samples were diagnosed using the TaqMan platform, which offers higher sensitivity and specificity compared to conventional microbiological methods, thereby improving estimates of pathogen-specific burden in community-based childhood diarrhea surveillance [8]. An earlier study using standardized conventional microbiology and multiplex polymerase chain reaction assay analyzed asymptomatic EAEC detections, alone or with co-pathogens, in the same population and stool samples. That study found that EAEC infection, especially at high pathogen loads, was associated with lower socioeconomic status, increased MPO levels, reduced NEO concentrations, and WLZ and WAZ—without significant impairment of intestinal function [7].

Enteric diseases disproportionately affect children in low-resource settings and are closely tied to poverty, which worsens both their incidence and severity [10]. Conditions linked to poverty—such as inadequate hygiene, poor sanitation, overcrowded housing, improper food storage, and restricted access to clean water—foster an environment conducive to the transmission of enteric pathogens. Additionally, close contact with domestic animals, which may serve as reservoirs for human pathogens, further elevates the risk of infection in these vulnerable populations [10, 22–24].

In our cohort, asymptomatic EAEC detections with high pathogen burdens were more prevalent among children exposed to socioeconomic risk factors such as low household income, low maternal education, inadequate sanitation, and unsafe drinking water. Indices of socioeconomic status, including a composite SES index based on household assets and the WAMI index (Water/Sanitation, Assets, Maternal education, and Income) appeared as significant predictors of EAEC coinfection and pathogen burden. These findings are consistent with prior research showing associations between enteric infections and lower socioeconomic indicators [7, 25].

Low maternal education was also identified as a significant risk factor for enteric infection. Mothers with lower educational attainment were more likely to have children infected with EAEC and at least two more pathogens. An earlier analysis in this cohort found maternal education to be a predictor of EAEC infections caused by distinct genomic strains and associated with linear growth faltering [10].

Interestingly, the proportion of breastfeeding days up to six months of age did not significantly differ between coinfected and uninfected children, contrary to earlier findings [7, 26]. Although breastfeeding in the first year of life is well-established as protective against enteric infections [9, 27, 28], in our study it did not appear to influence the occurrence of subclinical infections, including those involving EAEC. In contrast, the duration of antibiotic use appeared as a more relevant determinant, consistently associated with subclinical infections characterized by high pathogen burdens, with or without EAEC, and negating the protective effects of breastfeeding.

This finding raises concern, as it highlights a critical public health issue: the potential role of indiscriminate antibiotic use in driving antimicrobial resistance. Earlier data from this cohort showed frequent antibiotic use in early childhood, particularly for non-bloody diarrhea and non-specific respiratory symptoms [29]. Our findings align with those of Lima et al. [7], reinforcing the hypothesis that inappropriate antibiotic use increases enteropathogen burden and facilitates EAEC infection.

On the pathogenesis of subclinical coinfections, Lima et al. [7] earlier found associations between EAEC, high pathogen burden, and intestinal inflammation, with no corresponding increase in intestinal permeability. In contrast, our study showed elevated paracellular permeability in children with high pathogen burdens, irrespective of EAEC presence. Notably, this increase did not appear to reflect over epithelial barrier damage. EAEC infection, alone or in combination with one or 3 or more pathogens, was associated with elevated fecal NEO—but not MPO—suggesting immune activation consistent with intestinal inflammation.

Elevated fecal NEO in the absence of increased MPO may reflect macrophage activation driven by interferon-γ produced by Th1 cells, in the absence of acute neutrophil-mediated inflammation [30]. This pattern may be indicative of a non-destructive immune response or chronic low-grade immune activation, both characteristic of subclinical enteric infection and consistent with our bowel function findings.

More severe patterns of intestinal inflammation were reported in children with diarrhea from the same cohort [9], in whom EAEC infection was associated with elevated levels of both MPO and NEO, as well as greater intestinal dysfunction. In line with these findings, our results are consistent with transcriptomic analyses of duodenal biopsies from children with environmental enteropathy (EE), which revealed activation of the Janus kinase/signal transducer and activator of transcription (JAK/STAT) pathway, along with increased IL-18 and IL-17 signaling — hallmarks of cellular immune activation involving Th1 and Th17 responses [31]. Together, these results suggest that asymptomatic EAEC detection and similar enteric exposures can elicit targeted immune activation like EE without inducing overt intestinal damage.

Anthropometric analyses further revealed that EAEC infection and higher pathogen burdens were associated with poorer growth outcomes - reduced LAZ scores. The length-for-age delta was 0.44 units lower among infected children than among their pathogen-free counterparts. These results corroborate prior findings linking EAEC to impaired linear and ponderal growth [7, 9, 10]. Additionally, repeated detection of EAEC at three time points was associated with more pronounced growth deficits by age two, underscoring the cumulative impact of recurrent infections [9].

Interestingly, co-infection with EAEC and two more pathogens was associated with increased WLZ. The most often co-detected pathogens in this group were aEPEC, ETEC, adenovirus, and norovirus—all commonly linked to malnutrition [32, 33, 34–39]. This unexpected finding may reflect a potential catch-up growth effect, driven by two factors: the cumulative duration of antibiotic treatment, which could have mitigated the negative impact on growth [29], and a lower intensity of acute inflammation, as suggested by reduced MPO levels saw in this group.

The findings from this study highlight a significant and potentially lasting impact of early subclinical Enteroaggregative *Escherichia coli* (EAEC) infection on child neurodevelopment. Children exposed to EAEC alone during the first six months of life exhibited significantly lower BSID-III composite cognitive scores at both 15 and 24 months, as well as reduced language scores at 24 months, even after adjusting for socioeconomic and health-related covariates. These findings support the hypothesis that EAEC mono-infection may interfere with critical early neurodevelopmental processes, potentially through mechanisms involving gut-brain axis disruption, intestinal inflammation, and impaired nutrient absorption [10, 30, 40]. Prior studies have described how enteric infections, especially in early life, contribute to developmental delays by promoting chronic intestinal inflammation and environmental enteric dysfunction (EED), a subclinical condition that compromises nutrient uptake and systemic immunity [31, 40, 41].

Evidence from the MAL-ED cohort has demonstrated associations between EAEC burden and growth faltering [39], as well as increased intestinal inflammatory biomarkers indicative of EED [8]. Furthermore, elevated markers of EED, such as myeloperoxidase (MPO) and neopterin (NEO), have been associated with poorer cognitive outcomes [42], reinforcing the plausibility of an enteropathy-mediated pathway linking subclinical infection to neurodevelopmental impairment. Interestingly, the presence of co-pathogens did not show similar consistent associations, which may reflect complex interactions that modulate the host response or mask EAEC-specific effects. These findings underscore the importance of early identification and intervention strategies targeting EAEC, particularly in settings where children are exposed to high enteric pathogen burdens during the critical window of brain development. These mechanisms, together with prior MAL-ED findings on intestinal inflammation and growth faltering [8, 9], support the plausibility of an enteropathy-mediated pathway linking early asymptomatic enteric infection to long-term neurodevelopmental impairment.

EAEC was often detected alongside pathogens such as ETEC, aEPEC, norovirus, adenovirus, astrovirus, Campylobacter, tEPEC, Giardia, sapovirus, and rotavirus. ETEC, EPEC, norovirus, Campylobacter, and Giardia have independently been linked to linear growth faltering in children from low-resource settings [27, 32–39], suggesting additive or synergistic effects on growth impairment. In addition, *Shigella*, enteroaggregative *E coli, Campylobacter*, and *Giardia*, had a negative association with linear growth, which was significante during the first 2 to 5 years of life [39].

These growth deficits are associated, at least in part, by enteric inflammation. One study showed that intestinal inflammation—specifically elevated fecal MPO—was associated with systemic inflammation and growth hormone resistance in children with diarrhea [43]. Similarly, children with EE show intestinal inflammation, malabsorption, and increased permeability, leading to microbial translocation, systemic inflammation, and undernutrition [3, 44]. In our study, intestinal inflammation, reflected by elevated neopterin, may serve as a mechanistic link between EAEC infection, pathogen burden, and impaired growth.

Furthermore, a study by Rina et al. [10] found that the presence of EAEC virulence genes *aaiC* and *aatA* was associated with higher EE scores and increased intestinal inflammation. In this context, detection of at least one of these genes alongside high pathogen load may suggest EE in our cohort.

This study has several limitations. Monthly stool sampling may have missed intermittent pathogen shedding, potentially underestimating the frequency of co-infections. Additionally, infection duration was not assessed, which may influence outcomes. Incorporating biomarkers of microbial translocation and systemic inflammation would enhance characterization of asymptomatic EAEC detections and their contribution to EE and growth deficits. Despite these limitations, our findings provide novel evidence linking asymptomatic EAEC detection and co-infection in early childhood to intestinal inflammation, altered permeability, and impaired linear growth.

In conclusion, quantitative molecular diagnostics enabled the identification of robust associations between asymptomatic EAEC detection, pathogen burden, intestinal dysfunction, impaired growth, and neurodevelopment in early childhood. EAEC and high enteropathogen loads appeared as key contributors to intestinal dysfunction and potential environmental enteropathy. These results highlight the urgent need to improve access to clean water and sanitation and to restrict unnecessary antibiotic use in vulnerable populations, to mitigate subclinical pathogen exposure and its negative impacts on child growth and neurodevelopment (**Figure Summary Abstract**).

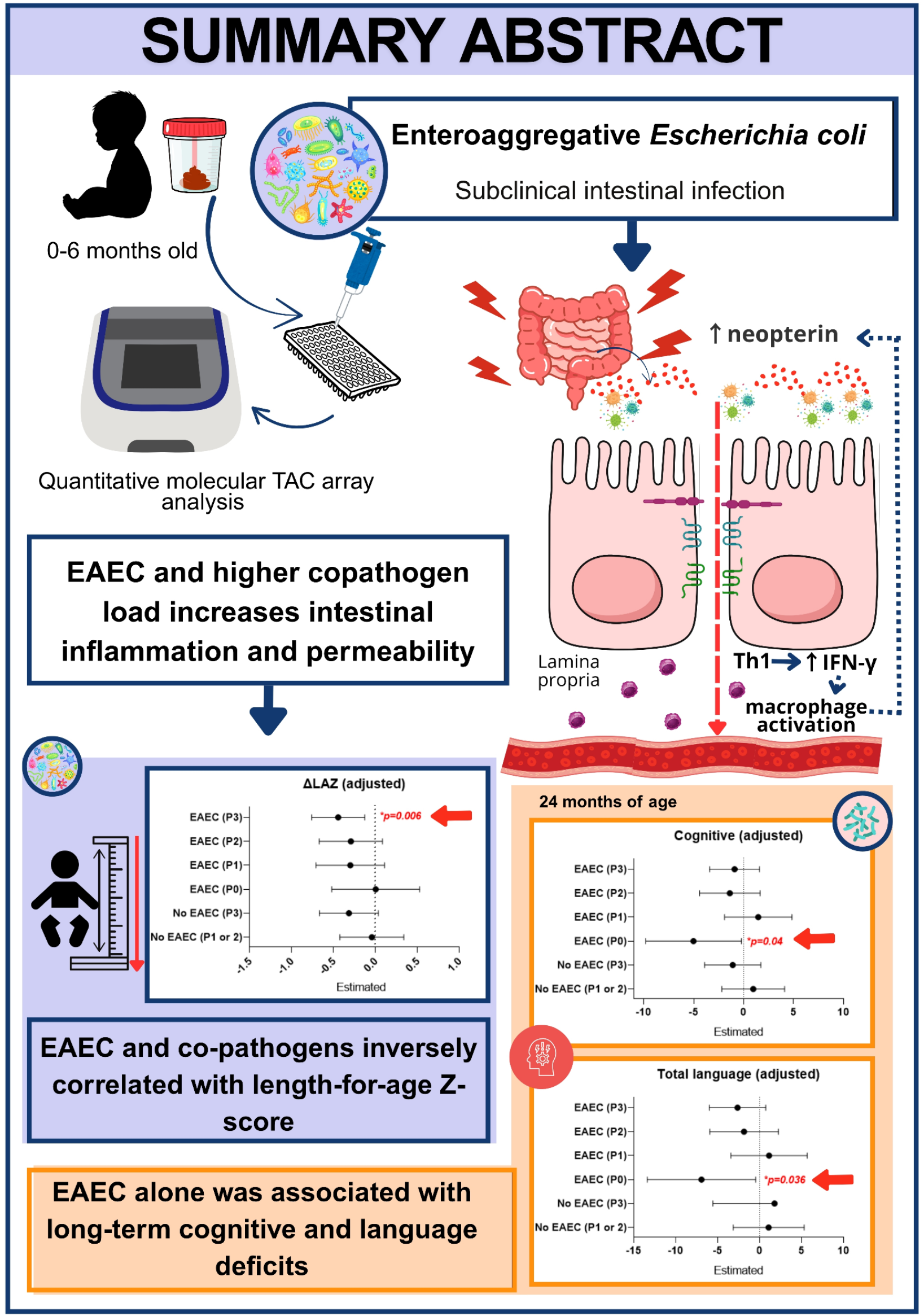

## Data Availability

All data produced in the present study are available upon reasonable request to the authors.

## Acknowledgements

The Etiology, Risk Factors and Interactions of Enteric Infections and Malnutrition and the Consequences for Child Health and Development Project (MAL-ED) is a collaborative project supported by the Bill & Melinda Gates Foundation (OPP1131125). The funding agencies had no responsibility for the design, analysis or writing of this article. All authors certify that they have no affiliations with or involvement in any organization or entity with any financial interest or non-financial interest in the subject matter or materials discussed in this manuscript.

## The authors had the following responsibilities

F.J.W.S and A.A.M.L.: outlined the research; V.L.S.C.U.: conducted the study; V.L.S.C.U., S.A.R., A.E.S.A, L.F.B.L., F.A.P.R, A.K.F.L., A.H. and A. A. M. L.: analyzed and interpreted data; V.L.S.C.U., S.A.R., J.Q.S.F., D.V.S.C. and A.A.M.L.: wrote the article and had final responsibility for the content; and all authors: read and approved the manuscript. The site PIs of the MAL-ED study and all co-authors have read and reviewed the manuscript.

## REFERENCES

1. Lima AAM, Moore SR, Barboza MS, et al. Persistent Diarrhea Signals a Critical Period of Increased Diarrhea Burdens and Nutritional Shortfalls: A Prospective Cohort Study among Children in Northeastern Brazil. J Infect Dis. 2000;181(5):1643–1651. DOI: 10.1086/315423.

2. Platts-Mills JA, Babji S, Bodhidatta L, et al. Pathogen-specific burdens of community diarrhea in developing countries: a multisite birth cohort study (MAL-ED). Lancet Glob Health. 2015;3(9):e564–e575. DOI: 10.1016/S2214-109X(15)00151-5.

3. Guerrant RL, DeBoer MD, Moore SR, et al. Biomarkers of environmental enteropathy, inflammation, stunting, and impaired growth in children in northeast Brazil. PLoS ONE. 2016;11(9):e0158772. DOI: 10.1371/journal.pone.0158772.

4. Adachi JA, Jiang ZD, Mathewson JJ, et al. Natural history of enteroaggregative and enterotoxigenic Escherichia coli infection among US travelers to Guadalajara, Mexico. J Infect Dis. 2002;185(11):1681–1683. DOI: 10.1086/340820.

5. Samie A, Obi CL, Barrett LJ, et al. Enteroaggregative Escherichia coli in Venda, South Africa: distribution of virulence-related genes by multiplex polymerase chain reaction in stool samples of HIV-positive and HIV-negative individuals and primary school children. Am J Trop Med Hyg 2007; 77:142–150.

6. Havt A, Lima IF, Lima AA, et al. Prevalência e perfil gênico de virulência de Escherichia coli enteroagregativa em crianças brasileiras desnutridas e nutridas. Diagn Microbiol Infect Dis. 2017;89(2):98–105. DOI: 10.1016/j.diagmicrobio.2017.06.014.

7. Lima AAM, Oriá RB, Soares AM, et al. Enteroaggregative Escherichia coli Subclinical Infection and Coinfections and Impaired Child Growth in the MAL-ED Cohort Study. J Pediatr Gastroenterol Nutr. 2018;66(2):325–333. DOI: 10.1097/MPG.0000000000001746.

8. Platts-Mills JA, Liu J, Rogawski ET, et al. Use of quantitative molecular diagnostic methods to assess the aetiology, burden, and clinical characteristics of diarrhea in children in low-resource settings: a reanalysis of the MAL-ED cohort study. Lancet Glob Health. 2018;6(12):e1309–e1318. DOI: 10.1016/S2214-109X(18)30349-8.

9. Rogawski ET, Guerrant RL, Havt A, et al. Epidemiology of enteroaggregative Escherichia coli infections and associated outcomes in the MAL-ED birth cohort. PLoS Negl Trop Dis. 2017;11(7):e0005798. DOI: 10.1371/journal.pntd.0005798.

10. Das R, Colston J, Seidman J, et al. Site specific incidence rate of virulence related genes of enteroaggregative Escherichia coli and association with enteric inflammation and growth in children. Sci Rep. 2021;11(1):23178. DOI: 10.1038/s41598-021-02626-z.

11. Richard, S. A., Mccormick, B. J. J., Miller, M. A., et al. Modeling environmental influences on child growth in the MAL-ED cohort study: opportunities and challenges. Clin Infect Dis. 2014; 59(Suppl 4): S255–S260. DOI: 10.1093/cid/ciu436

12. Murray-Kolb LE, Rasmussen ZA, et al. The MAL-ED Cohort Study: Methods and Lessons Learned When Assessing Early Child Development and Caregiving Mediators in Infants and Young Children in Low- and Middle-Income Countries, Clin Infect Dis. 2014; 59(Suppl 4): S261–S272. DOI: 10.1093/cid/ciu437

13. Houpt E, Gratz J, Kosek K, et al. The MAL-ED Network Investigators, Microbiologic Methods Utilized in the MAL-ED Cohort Study. Clin Infect Dis. 2014; 59(Suppl 4): S225–S232, 10.1093/cid/ciu413

14. World Health Organization Multicentre Growth Reference Study Group. WHO child growth standards based on length/height, weight and age. Acta Paediatr Suppl 2006;450:76–85. DOI: 10.1111/j.1651-2227.2006.tb02378.x.

15. Kosek M, Haque R, Lima A, et al. Fecal Markers of Intestinal Inflammation and Permeability Associated with the Subsequent Acquisition of Linear Growth Deficits in Infants. ASTMH. 2013;88(2):390–396. doi:10.4269/ajtmh.2012.12-0549.

16. MAL-ED Network Investigators. The MAL-ED study: a multinational and multidisciplinary approach to understand the relationship between enteric pathogens, malnutrition, gut physiology, physical growth, cognitive development, and immune responses in infants and children up to 2 years of age in resource-poor environments. Clin Infect Dis. 2014;59(Suppl 4):S193–S206. DOI: 10.1093/cid/ciu653.

17. Bayley N. Bayley Scales of Infant and Toddler Development. San Antonio, TX: The Psychological Corporation, 2006.

18. Fagundes NU, Affonso SIC. Escherichia coli infections and malnutrition. Lancet. 2000 Dec;356 Suppl:s27. DOI: 10.1016/S0140-6736(00)92013-0.

19. Mondal D, Petri WA, Sack RB, et al. Attribution of malnutrition to cause-specific diarrheal illness: evidence from a prospective study of preschool children in Mirpur, Dhaka, Bangladesh. Am J Trop Med Hyg. 2018;80(5):824–826. DOI: 10.4269/ajtmh.2009.80.824.

20. Dias RCB, Dias VF, Pinheiro SR, et al. Analysis of the virulence profile and phenotypic features of typical and atypical enteroaggregative Escherichia coli (EAEC) isolated from diarrheal patients in Brazil. Front Cell Infect Microbiol. 2020;10:144. DOI: 10.3389/fcimb.2020.00144.

21. Modgil V, Ghosh A, Mandal J. Analysis of the virulence and inflammatory markers elicited by enteroaggregative Escherichia coli isolated from clinical and non-clinical sources in an experimental infection model, India. Microbiol Res. 2022;13(4):882–897. DOI: 10.3390/microbiolres13040065.

22. Null C, Stewart CP, Pickering AJ, et al. Effects of water quality, sanitation, handwashing, and nutritional interventions on diarrhoea and child growth in rural Kenya: a cluster-randomised controlled trial. Lancet Glob Health. 2018;6(3):e316–e329. DOI: 10.1016/S2214-109X(18)30005-6.

23. Luby SP, Rahman M, Arnold BF, et al. Effects of water quality, sanitation, handwashing, and nutritional interventions on diarrhoea and child growth in rural Bangladesh: a cluster randomised controlled trial. Lancet Glob Health. 2018;6(3):e302–e315. DOI: 10.1016/S2214-109X(17)30490-4.

24. Magalhães Lmvc, Lima NL, Lima AAM, et al. Quantifying lactulose and mannitol using LC-MS/MS in a clinical study of children with environmental enteric disease. Braz J Med Biol Res. 2025;58:e12345. DOI: 10.1590/1414-431X2025e12345.

25. Colston JM, Saboorian R, Delahoy MJ, et al. Associations between household-level exposures and all-cause diarrhea and pathogen-specific enteric infections in children enrolled in five sentinel surveillance studies. Int J Environ Res Public Health. 2020;17(21):8078. DOI: 10.3390/ijerph17218078.

26. McCormick BJJ, Lee GO, Seidman JC, et al. Full breastfeeding protection against common enteric bacteria and viruses: results from the MAL-ED cohort study. Am J Clin Nutr. 2022;115(3):759–769. DOI: 10.1093/ajcn/nqab377.

27. Rogawski ET, Bartelt LA, Seidman JC, et al. Determinants and impact of Giardia infection in the first 2 years of life in the MAL-ED birth cohort. J Pediatric Infect Dis Soc 2017c; x:1–8. DOI: 10.1093/jpids/piw082.

28. Davisse-Paturet C, Adel-Patient K, Pierson J, et al. Breastfeeding Status and Duration and Infections, Hospitalizations for Infections, and Antibiotic Use in the First Two Years of Life in the ELFE Cohort. Nutrients. 2019;11(7):1607. DOI: 10.3390/nu11071607.

29. Rogawski ET, Platts-Mills JA, Seidman JC, et al. Use of antibiotics in children younger than two years in eight countries: a prospective cohort study. Bull World Health Organ. 2017;95(1):49–61. DOI: 10.2471/BLT.

30. Murr C, Widner B, Wirleitner B, Fuchs D. Neopterin as a marker for immune system activation. Curr Drug Metab. 2002;3(2):175–187. DOI: 10.2174/1389200024605082.

31. Marie C, Das S, Coomes D, et al. Duodenal transcriptomics demonstrates signatures of tissue inflammation and immune cell infiltration in children with environmental enteric dysfunction across global centers. *Am J Clin Nutr*. 2024;120 Suppl 1:S51–64. DOI: 10.1016/j.ajcnut.2024.02.023

32. Gondim RDG, Carvalho-Costa FA, Oliveira DS, et al. Genetic diversity of norovirus infections, co-infections, and undernutrition in children from Brazilian semiarid region. J Pediatr Gastroenterol Nutr. 2018;67(4):e82–e86. DOI: 10.1097/MPG.0000000000002043.

33. Do Nascimento Veras H, Oriá RB, Lima AA, et al. Campylobacter jejuni virulence genes and immune-inflammatory biomarkers association with growth impairment in children from Northeastern Brazil. Eur J Clin Microbiol Infect Dis. 2018;37(10):2011–2020. DOI: 10.1007/s10096-018-3337-0.

34. Das R, Colston J, Seidman J, et al. Incidence of genomic subtypes of EPEC and association with inflammation and growth. Sci Rep. 2022;12(1):5724. DOI: 10.1038/s41598-022-09730-8.

35. Rogawski McQuade ET, Platts-Mills JA, Gratz J, et al. Epidemiology of Shigella species and serotypes in children: a retrospective substudy of the MAL-ED observational birth cohort study. Lancet Glob Health 2025. Online First: 26 Mar. DOI: 10.2139/ssrn.4896053.

36. Schiaffino F, Paredes Olortegui M, Peñataro Yori P, et al. The epidemiology and impact of persistent Campylobacter infections on childhood growth among children 0–24 months of age in resource-limited settings. EClinicalMedicine 2024; 76:102841. DOI: 10.1016/j.eclinm.2024.102841.

37. Gazi MA, Jahan N, Mahfuz M, et al. Infection with E. coli pathotypes and association with gut enteropathy and nutritional status in Bangladesh. Front Cell Infect Microbiol. 2022; 12:901324. DOI: 10.3389/fcimb.2022.901324.

38. Pajuelo MJ, Del Canto F, Chauca J, et al. Epidemiology of enterotoxigenic Escherichia coli and impact on child growth in Lima, Peru. Front Public Health. 2024;12:1332319. DOI: 10.3389/fpubh.2024.1332319.

39. Rogawski ET, Liu J, Platts-Mills JA, et al. Use of quantitative molecular diagnostic methods to investigate the effect of enteropathogen infections on linear growth in children in low-resource settings: longitudinal analysis of results from the MAL-ED cohort study. Lancet Glob Health 2018. DOI: 10.1016/S2214-109X(18)30351-6.

40. Guerrant RL, DeBoer MD, Moore SR, Scharf RJ, Lima AA. The impoverished gut—a triple burden of diarrhoea, stunting and chronic disease. Nat Rev Gastroenterol Hepatol. 2013;10(4):220–9. doi:10.1038/nrgastro.2012.239. PMID: 23380764.

41. Keusch GT, Denno DM, Black RE, Duggan C, Guerrant RL, Lavery JV, Tarr PI, Ward HD, Nataro JP, Ryan ET, Houpt ER. Environmental enteric dysfunction: pathogenesis, diagnosis, and clinical consequences. Clin Infect Dis. 2014;59(Suppl\_4)\:S207–12. doi:10.1093/cid/ciu485. PMID: 25305293.

42. Scharf RJ, Rogawski ET, Murray-Kolb LE, Maphula A, Maphula N, Svensen E, et al. Early childhood growth and cognitive outcomes: findings from the MAL-ED study. Matern Child Nutr. 2018;14(3):e12584. doi: 10.1111/mcn.12584.

43. DeBoer MD, Scharf RJ, Leite AM, et al. Systemic inflammation, growth factors, and linear growth in the setting of infection and malnutrition. Nutrition. 2017; 33:248–253. doi: 10.1016/j.nut.2016.06.013.

44. Bartelt LA, Bolick DT, Guerrant RL. Disentangling Microbial Mediators of Malnutrition: Modeling Environmental Enteric Dysfunction. Cell Mol Gastroenterol Hepatol. 2019;7(3):692–707. DOI: 10.1016/j.jcmgh.2018.12.006.

