## Supplementary Material for "Impact of Asymptomatic Enteroaggregative *Escherichia coli* Infection and Co-Pathogen Burden on Intestinal Barrier Function, Linear Growth, and Cognitive Development in Early Childhood: Insights from a Birth Cohort Study"

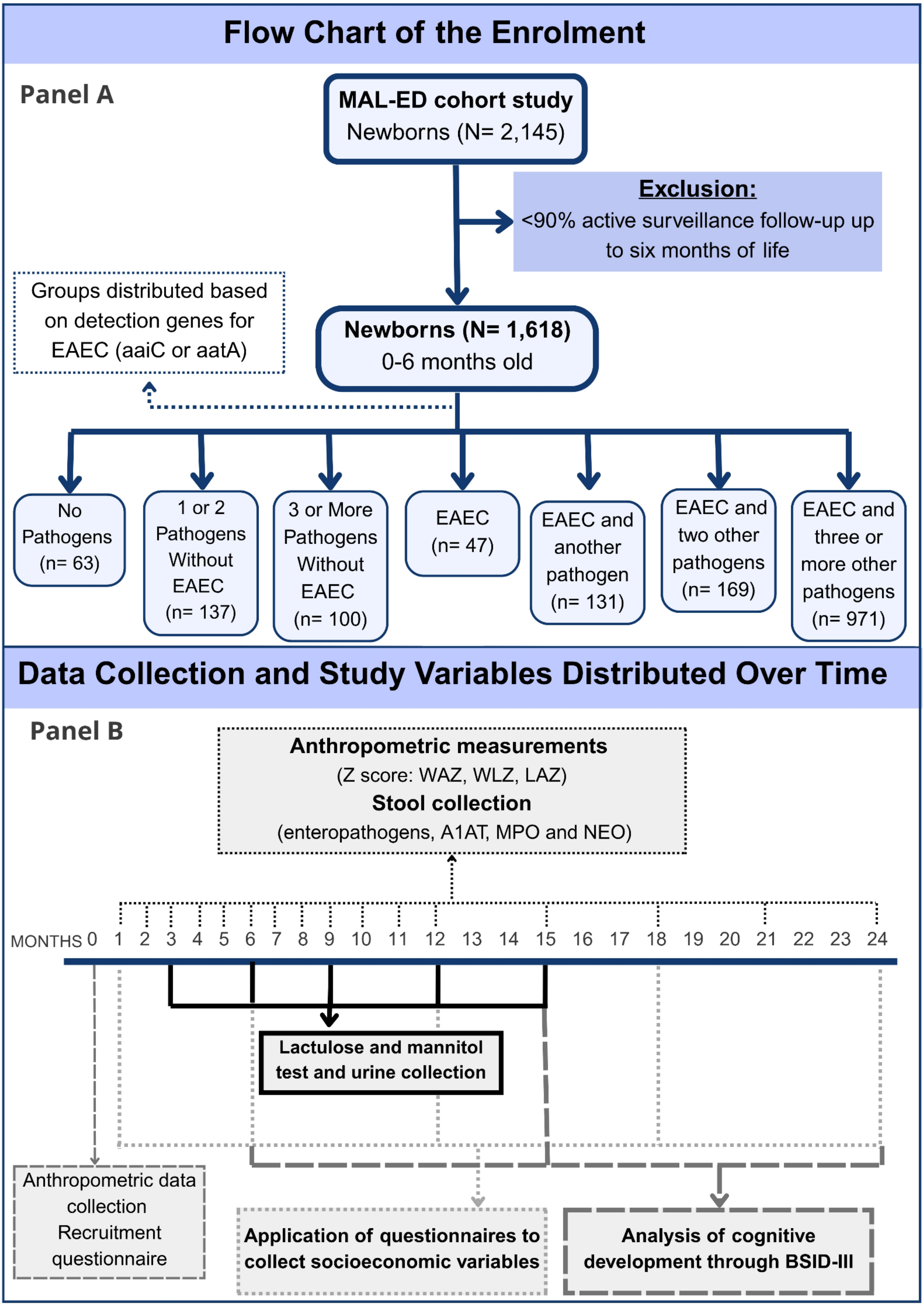

**Figure 1** – Flowchart of recruitment and follow-up procedures in the MAL-ED cohort study, panel A. A total of 2,145 newborns were enrolled across eight study sites between November 2009 and February 2012. Infants with less than 90% active surveillance follow-up through the first six months of life were excluded, yielding a final analytical sample of 1,659 infants. These participants were stratified into seven groups based on the presence of the aaiC or aatA genes (used for EAEC detection) and the number of copathogens detected (categorized as 1, 2, or ≥3). Panel B, Data collection included: baseline anthropometry and a recruitment questionnaire at birth; subsequent anthropometric assessments (Z-scores: WAZ, WLZ, LAZ); stool sample collection for enteropathogen analysis, A1AT, MPO, and NEO quantification; questionnaires on socioeconomic variables; the Bayley Scales of Infant and Toddler Development, Third Edition (BSID-III) were used to assess cognitive development; and the lactulose-mannitol (LM) test with corresponding urine collection. The timing of each procedure is detailed above.

**Table Suppl. 1** – Adjusted and unadjusted linear regression model to evaluate biomarkers of intestinal function and inflammation in children aged 0 to 6 months with or without subclinical infection.

| | | No EAEC (P1 or P2) | | No EAEC (P3) | | EAEC (P0) | | EAEC (P1) | | EAEC (P2) | | EAEC (P3) | |
|---|---|---|---|---|---|---|---|---|---|---|---|---|---|
| **Model** | **Outcome variable** | **β (95% CI)** | ***p*** | **β (95% CI)** | ***p*** | **β (95% CI)** | ***p*** | **β (95% CI)** | ***p*** | **β (95% CI)** | ***p*** | **β (95% CI)** | ***p*** |
| **Unadjusted** | Z escore % Lactulose | -0.05 (-0.47, 0.36) | 0.797 | **0.40 (0.01, 0.79)** | **0.042** | 0.099 (-0.45, 0.65) | 0.726 | 0.32 (-0.12, 0.77) | 0.154 | 0.24 (-0.17, 0.65) | 0.257 | 0.31 (-0.03, 0.65) | 0.075 |
| | Z escore % Mannitol | 0.0007 (-0.3, 0.3) | 0.996 | 0.05 (-0.22, 0.32) | 0.717 | -0.22 (-0.6, 0.16) | 0.268 | 0.0001 (-0.31, 0.31) | 1.000 | -0.11 (-0.4, 0.18) | 0.451 | 0.02 (-0.21, 0.26) | 0.843 |
| | Z escore L/M | -0.075 (-0.33, 0.18) | 0.568 | 0.17 (-0.06, 0.41) | 0.145 | 0.26 (-0.08, 0.6) | 0.133 | 0.15 (-0.12, 0.42) | 0.277 | 0.21 (-0.04, 0.5) | 0.097 | 0.11 (-0.09, 0.32) | 0.280 |
| | MPO | 1817.7 (-1487, 5122) | 0.281 | 474.2 (-2567, 3515) | 0.760 | 36.3 (-4196, 4268) | 0.987 | 2840.7 (-629, 6310) | 0.108 | 430.8 (-2804, 3666) | 0.794 | **3509.4 (788, 6231)** | **0.012** |
| | NEO | 332 (-377, 1040) | 0.359 | 230 (-422, 882) | 0.490 | 517 (-390, 1425) | 0.264 | 553 (-191, 1297) | 0.145 | 412 (-282, 1106) | 0.244 | 175 (-409, 759) | 0.556 |
| | A1AT | 0.04 (-0.09, 0.18) | 0.502 | 0.06 (-0.06, 0.18) | 0.316 | 0.13 (-0.04, 0.3) | 0.124 | 0.05 (-0.09, 0,2) | 0.459 | -0.04 (-0.17, 0.09) | 0.552 | -0.004 (-0.11, 0.1) | 0.942 |
| **Adjusted** | Z escore % Lactulose | -0.003 (-0.44, 0.43) | 0.988 | **0.44 (0.046, 0.84)** | **0.029** | 0.12 (-0.45, 0.7) | 0.673 | 0.33 (-0.13, 0.78) | 0.160 | 0.34 (-0.085, 0.77) | 0.117 | **0.41 (0.058, 0.77)** | **0.023** |
| | Z escore % Mannitol | 0.017 (-0.28, 0.32) | 0.910 | 0.11 (-0.16, 0.39) | 0.431 | -0.22 (-0.61, 0.19) | 0.289 | 0.087 (-0.23, 0.40) | 0.586 | 0.044 (-0.25, 0.34) | 0.772 | 0.10 (-0.14, 0.35) | 0.422 |
| | Z escore L/M | -0.056 (-0.33, 0.21) | 0.683 | 0.16 (-0.09, 0.40) | 0.216 | 0.28 (-0.08, 0.63) | 0.128 | 0.084 (-0.20, 0.37) | 0.563 | 0.15 (-0.11, 0.42) | 0.251 | 0.12 (-0.10, 0.34) | 0.304 |
| | MPO | 535.24 (-2798.2, 3868.7) | 0.753 | -769.78 (-3812.7, 2273.1) | 0.620 | -293.03 (-4608.6, 4022.5) | 0.894 | 2589.41 (-888.8, 6067.6) | 0.144 | -1.65 (-3261.4, 3258.1) | 0.999 | 693.80 (-2069.8, 3457.4) | 0.622 |
| | NEO | 486.5 (-186.4, 1159.27) | 0.156 | 543.5 (-70.7, 1157.70) | 0.083 | 595.7 (-275.4, 1466.72) | 0.180 | 1016.4 (314.4, 1718.46) | **0.005** | 381.5 (-276.4, 1039.46) | 0.256 | **687.5 (129.7, 1245.31)** | **0.016** |
| | A1AT | 0.059 (-0.08, 0.2) | 0.404 | 0.067 (-0.06, 0.2) | 0.295 | 0.072 (-0.11, 0.25) | 0.430 | 0.065 (-0.08, 0.21) | 0.374 | 0.003 (-0.13, 0.14) | 0.963 | 0.005 (-0.11, 0.12) | 0.926 |

MPO: Myeloperoxidase, ng/mL; NEO: Neopterin, nmol/L; A1AT: Alpha-1-antitrypsin, mg/g; β: β-estimate; CI: confidence interval; p: statistical value.

% Lactulose, % mannitol, and lactulose:mannitol ratio Z scores assigned median values over 3- and 6-months values to age cumulative interval 0 to 6 months.

Myeloperoxidase, neopterin, and alpha-1-antitrypsin assigned median values over the cumulative interval 0 to 6 months.

All groups were compared to the non-pathogen group.

Multivariate linear regression had the following covariates as adjustment: location, household food insecurity, maternal education, socioeconomic status (WAMI index), and the percentage of days with antibiotic use.

**Table Suppl. 2** – Adjusted and unadjusted linear regression model to evaluate anthropometric scores (delta LAZ, WAZ, WLZ 0-6m) in children aged 0 to 6 months with or without subclinical infection.

| | | No EAEC (P1 or P2) | | No EAEC (P3) | | EAEC (P0) | | EAEC (P1) | | EAEC (P2) | | EAEC (P3) | |
|---|---|---|---|---|---|---|---|---|---|---|---|---|---|
| **Model** | **Outcome variable** | **β (95% CI)** | ***p*** | **β (95% CI)** | ***p*** | **β (95% CI)** | ***p*** | **β (95% CI)** | ***p*** | **β (95% CI)** | ***p*** | **β (95% CI)** | ***p*** |
| **Unadjusted** | ΔLAZ | -0.17 (-0.53, 0.2) | 0.372 | **-0.45 (-0.8, -0.11)** | **0.009** | -0.08 (-0.57, 0.41) | 0.747 | **-0.42 (-0.81, -0.02)** | **0.037** | **-0.42 (-0.78, -0.06)** | **0.023** | **-0.72 (-1.02, -0.42)** | **<.001** |
| | ΔWAZ | 0.29 (-0.12, 0.7) | 0.164 | -0.05 (-0.44, 0.33) | 0.775 | 0.19 (-0.36, 0.75) | 0.487 | -0.14 (-0.6, 0.3) | 0.519 | 0.16 (-0.25, 0.57) | 0.445 | **-0.43 (-0.77, -0.1)** | **0.011** |
| | ΔWLZ | **0.55 (0.02, 1.1)** | **0.040** | 0.36 (-0.13, 0.85) | 0.151 | 0.35 (-0.35, 1.05) | 0.331 | 0.16 (-0.4, 0.73) | 0.578 | **0.57 (0.05, 1.1)** | **0.032** | 0.035 (-0.4, 0.5) | 0.874 |
| **Adjusted** | ΔLAZ | -0.04 (-0.42, 0.34) | 0.838 | -0.31 (-0.66, 0.04) | 0.081 | 0.006 (-0.52, 0.53) | 0.981 | -0.3 (-0.7, 0.11) | 0.154 | -0.3 (-0.67, 0.09) | 0.131 | **-0.44 (-0.76, -0.12)** | **0.006** |
| | ΔWAZ | 0.33 (-0.08, 0.75) | 0.115 | 0.01 (-0.4, 0.4) | 0.946 | 0.22 (-0.35, 0.8) | 0.439 | -0.003 (-0.45, 0.44) | 0.988 | 0.3 (-0.12, 0.71) | 0.163 | -0.14 (-0.49, 0.2) | 0.419 |
| | ΔWLZ | 0.48 (-0.06, 1.02) | 0.084 | 0.34 (-0.16, 0.84) | 0.184 | 0.27 (-0.48, 1.01) | 0.480 | 0.26 (-0.32, 0.83) | 0.386 | **0.63 (0.1, 1.2)** | **0.021** | 0.2 (-0.25, 0.65) | 0.390 |

Enteroaggregative *Escherichia coli* (EAEC) with or none (P0), 1 (P1), 2 (P2), and 3 (P3) or more any other pathogens. All groups were compared to the non-pathogen group. β: β-estimate; CI: confidence interval; p: statistical value. Multivariate linear regression had the following covariates as adjustment: location, household food insecurity, maternal education, socioeconomic status (WAMI index), and the percentage of days with antibiotic use.

ΔLAZ: delta length-for-age Z-score; ΔWAZ: delta weight-for-age Z-score; ΔWLZ: delta weight-for-length Z-score. The delta scores correspond to the values at 0 and 6 months of age.

**Table Suppl. 3 –** Unadjusted and adjusted linear regression model to evaluate Baeley scales (cognitive scale, total language and motor scores, and social-emotional scale) in children aged 15 and 24 months with or without subclinical infection.

| | | No EAEC (P1 or P2) | | No EAEC (P3) | | EAEC (P0) | | EAEC (P1) | | EAEC (P2) | | EAEC (P3) | |
|---|---|---|---|---|---|---|---|---|---|---|---|---|---|
| **Model** | | **β (95% CI)** | ***p*** | **β (95% CI)** | ***p*** | **β (95% CI)** | ***p*** | **β (95% CI)** | ***p*** | **β (95% CI)** | ***p*** | **β (95% CI)** | ***p*** |
| **Bayley scales at 15 months:** | | | | | | | | | | | | | |
| Unadjusted | Cognitive | -0.20 (-2.02, 1.63) | 0.832 | -0.18 (-1.84, 1.48) | 0.833 | **-2.81 (-5.13, -0.50)** | **0.017** | 0.55 (-1.35, 2.44) | 0.571 | -0.56 (-2.33, 1.22) | 0.538 | -0.85 (-2.32, 0.624) | 0.259 |
| | Total language | 0.80 (-1.47, 3.06) | 0.492 | -1.34 (-3.40, 0.73) | 0.204 | -1.23 (-4.10, 1.66) | 0.404 | -0.77 (-3.12, 1.59) | 0.522 | -0.54 (-2.7, 1.66) | 0.629 | -1.06 (-2.89, 0.77) | 0.257 |
| | Total motor | 0.44 (-1.81, 2.69) | 0.702 | -0.10 (-2.15, 1.95) | 0.923 | -2.38 (-5.24, 0.47) | 0.102 | 0.37 (-1.97, 2.70) | 0.758 | -0.20 (-2.38, 1.98) | 0.858 | -0.70 (-2.51, 1.12) | 0.453 |
| | Social-emotional | 1.19 (-4.16, 6.53) | 0.664 | -4.83 (-9.70, 0.024) | 0.051 | 0.44 (-6.34, 7.22) | 0.899 | -2.41 (-7.96, 3.14) | 0.395 | **-5.66 (-10.85, -0.48)** | **0.032** | **-5.73 (-10.04, -1.42)** | **0.009** |
| Adjusted | Cognitive | 0.09 (-1.72, 1.89) | 0.925 | 0.55 (-1.08, 2.17) | 0.509 | **-2.82 (-5.15, -0.48)** | **0.018** | 1.42 (-0.45, 3.28) | 0.136 | 0.14 (-1.62, 1.89) | 0.880 | 0.10 (-1.36, 1.56) | 0.509 |
| | Total language | 0.53 (-1.69, 2.74) | 0.641 | -0.77 (-2.76, 1.22) | 0.448 | -1.61 (-4.46, 1.25) | 0.270 | 0.15 (-2.14, 2.43) | 0.901 | 0.07 (-2.08, 2.22) | 0.948 | -0.51 (-2.30, 1.27) | 0.573 |
| | Total motor | 0.27 (-1.89, 2.42) | 0.807 | 0.76 (-1.18, 2.70) | 0.441 | -2.43 (-5.21, 0.35) | 0.087 | 1.57 (-0.66, 3.79) | 0.167 | 0.89 (-1.20, 2.98) | 0.405 | 0.64 (-1.10, 2.38) | 0.471 |
| | Social-emotional | 1.71 (-3.17, 6.58) | 0.492 | -2.28 (-6.66, 2.10) | 0.307 | 2.74 (-3.54, 9.03) | 0.392 | 0.73 (-4.29, 5.76) | 0.775 | -2.14 (-6.87, 2.58) | 0.374 | -0.90 (-4.83, 3.04) | 0.654 |
| **Bayley scales at 24 months:** | | | | | | | | | | | | | |
| Unadjusted | Cognitive | 2.09 (-1.26, 5.43) | 0.221 | -0.69 (-3.74, 2.37) | 0.659 | -1.37 (-6.16, 3.42) | 0.575 | 0.93 (-2.66, 4.53) | 0.611 | -0.52 (-3.74, 2.71) | 0.753 | 0.12 (-2.56, 2.81) | 0.928 |
| | Total language | 2.36 (-2.35, 7.07) | 0.326 | -2.49 (-6.80, 1.81) | 0.256 | -2.98 (-9.73, 3.78) | 0.387 | -0.71 (-5.77, 4.36) | 0.785 | -1.17 (-5.71, 3.37) | 0.613 | -3.10 (-6.88, 0.68) | 0.108 |
| | Total motor | 1.55 (-1.98, 5.08) | 0.389 | -0.65 (-3.88, 2.57) | 0.691 | 0.51 (-4.55, 5.58) | 0.843 | -0.79 (-4.59, 3.01) | 0.683 | 0.45 (-3.86, 2.95) | 0.794 | 1.71 (-4.54, 1.13) | 0.237 |
| | Social-emotional | 6.89 (-2.03, 15.82) | 0.130 | -5.73 (-13.88, 2.43) | 0.168 | 5.98 (-6.82, 18.77) | 0.359 | 1.05 (-8.55, 10.64) | 0.830 | -1.13 (-9.73, 7.48) | 0.798 | **-8.10 (-15.26, -0.94)** | **0.027** |
| Adjusted | Cognitive | 0.96 (-2.18, 4.10) | 0.550 | -1.09 (-3.90, 1.72) | 0.447 | **-5.02 (-9.81, -0.22)** | **0.040** | 1.47 (-1.91, 4.85) | 0.393 | -1.38 (-4.42, 1.65) | 0.371 | -0.90 (-3.40, 1.59) | 0.477 |
| | Total language | 1.08 (-3.14, 5.31) | 0.615 | -1.78 (-5.56, 2.01) | 0.357 | **-6.92 (-13.38, -0.47)** | **0.036** | 1.13 (-3.42, 5.68) | 0.625 | -1.85 (-5.94, 2.24) | 0.374 | -2.63 (-5.99, 0.73) | 0.125 |
| | Total motor | 0.30 (-2.95, 3.55) | 0.856 | -0.36 (-3.27, 2.55) | 0.808 | -0.34 (-5.30, 4.61) | 0.892 | 0.20 (-3.29, 3.69) | 0.910 | -0.85 (-3.99, 2.29) | 0.595 | -1.49 (-4.07, 1.08) | 0.256 |
| | Social-emotional | 4.68 (-3.73, 13.08) | 0.275 | -3.12 (-10.65, 4.40) | 0.415 | 1.32 (-11.52, 14.14) | 0.841 | 5.08 (-3.97, 14.12) | 0.271 | -2.18 (-10.30, 5.94) | 0.599 | -4.65 (-11.33, 2.03) | 0.172 |

All groups were compared to the non-pathogen group. Enteroaggregative *Escherichia coli* (EAEC) with or none (P0), 1 (P1), 2 (P2), and 3 (P3) or more any other pathogens.
β: β-estimate; CI: confidence interval; p: statistical value. Multivariate linear regression had the following covariates as adjustment: location, household food insecurity, maternal education, socioeconomic status (WAMI index), and the percentage of days with antibiotic use.
The Bayley Scales of Infant included the following domains assessed: 1. Cognitive Scale assesses sensorimotor exploration, attention, memory, problem-solving, object exploration and pattern recognition; 2. Total Language Score included two subtests: (a) Receptive Language: understanding of words, sentences, commands and (b) Expressive Language: vocalizations and production of words/phrases; 3. Total Motor Score included two subdomains: (a) Fine Motor: eye-hand coordination, grasping, object manipulation and (b) Gross Motor: head control, sitting, crawling, walking and running; 4. Social-Emotional Scale was based on parent report (questionnaire): (a) Emotional regulation; (b) Social engagement; (c) Relationship with caregivers; and (d) Responses to stressors.
